# Newborn care knowledge and practices among care givers of new-born babies attending a regional referral hospital in Southwestern Uganda

**DOI:** 10.1101/2023.09.28.23296303

**Authors:** Dorah Nampijja, Kyoyagala Stella, Najjingo Elizabeth, Najjuma N. Josephine, Byamukama Onesmus, Kyasimire Lydia, Kabakyenga Jerome, Elias Kumbakumba

## Abstract

**Introduction:** A child born in developing countries has a 10 times higher mortality risk compared to one born in developed countries. Uganda still struggles with a high neonatal mortality rate at 27/1000 live births. Majority of these death occur in the community when children are under the sole care of their parents and guardian. Lack of knowledge in new born care, inappropriate new born care practices are some of the contributors to neonatal mortality in Uganda. Little is known about parent/caregivers’ knowledge, practices and what influences these practices while caring for the new borns. We systematically studied and documented newborn care knowledge, practices and associated factors among parents and care givers.

**Objective:** To assess new born care knowledge, practices and associated factors among parents and care givers attending MRRH

**Methods:** We carried out a quantitative cross section methods study among caregivers of children from birth to six weeks of life attending a regional referral hospital in south western Uganda. Using pretested structured questionnaires, data was collected about care givers’ new born care knowledge, practices and the associated factors. Data analysis was done using Stata version 17.0

**Results:** We interviewed 370 caregivers, majority of whom were the biological mothers at 86%. Mean age was 26 years, 14% were unemployed and 74% had monthly earning below the poverty line. Mothers had a high antenatal care attendance of 97.6% and 96.2% of the deliveries were at a health facility Care givers had variant knowledge of essential newborn care with associated incorrect practices. Majority (84.6%) of the respondents reported obliviousness to putting anything in the babies’ eyes at birth, however, breastmilk, water and saliva were reportedly put in the babies’ eyes at birth by some caregivers. Hand washing was not practiced at all in 16.2% of the caregivers before handling the newborn. About 7.4% of the new borns received a bath within 24 hours of delivery and 19% reported use of herbs. Caregivers practiced adequate thermal care 87%. Cord care practices were inappropriate in 36.5%. Only 21% of the respondents reported initiation of breast feeding within 1 hour of birth, Prelacteal feeds were given by 37.6% of the care givers, water being the commonest prelacteal feed followed by cow’s milk at 40.4 and 18.4% respectively. Majority of the respondents had below average knowledge about danger signs in the newborn where 63% and mean score for knowledge about danger signs was 44%. Caretaker’s age and relationship with the newborn were found to have a statistically significant associated to knowledge of danger signs in the newborn baby.

**Conclusion:** There are numerous incorrect practice in the essential new born care and low knowledge and awareness of danger signs among caregivers of newborn babies. There is high health center deliveries and antenatal care attendance among the respondents could be used as an opportunity to increase caregiver awareness about the inappropriate practices in essential newborn care and the danger signs in a newborn.

## Introduction

Globally, more than 8 million children die before their 5^th^ birthday (1), and neonatal deaths accounts for half of all under five mortality(2) In 2018, 2.8 million children died before one month of age worldwide. A child born in developing countries has a 10 times higher mortality risk compared to one born in developed countries(3, 4) Sub Saharan Africa faces the biggest burden with 1.2 million neonatal deaths every year(5). SSA has the highest neonatal mortality rate in the sustainable development goal regions at 28/1000 live birth. Uganda’s neonatal mortality rate has stagnated at 27/1000 live birth over the past 10 years(6), and large number of new born deaths occur in the communities under the sole care of their parents and care takers(7-9).

WHO recommends essential newborn care including early initiation of breast feeding, keeping babies warm, recognition of neonatal danger signs and cord care, among others, as crucial in new born survival(10). Most parents care for their new born babies with knowledge acquired from friends and family. This knowledge is often wrong and inappropriate. Once a baby is born in the community or discharged from a health facility, his/her care is entirely in the hands of the parents (often the mother) and the care givers. Lack of knowledge in new born care, inappropriate new born care practices, low health center deliveries among others contribute to the high neonatal deaths (9, 11).

Understanding new born care knowledge and practices among parents and care givers is crucial to improve survival among new born children. Parent centered new born care packages have been studied in some countries and have proved beneficial in new born outcomes(12). There is no available parent centered programs for new born care in Uganda and little is known about what parents /caregivers know or do and what influences their actions while they care for their new born babies at home. We set out to assess new born care knowledge, practices and associated factors among parents and care givers to newborn babies at Mbarara Regional Referral Hospital. This information will help to design feasible and acceptable high impact new born care packages and set a baseline for health education for parents and care givers of new born children.

## Methods

### Study area

We conducted a cross sectional, quantitative hospital based study from November 2022 to February 2023 at a regional referral hospital in South western Uganda with a catchment population of 3 million people with a bed capacity of 494 beds. The hospital also serves as a teaching hospital for Mbarara University of Science and Technology. The hospital offers a number of services including antenatal care, delivery and obstetric care, paediatric care including a newborn and premature care unit. It serves as a referral hospital in South-western region for more than 15 districts with a catchment population of 10 million people. The hospital also receives patients from the neighboring regional referral hospitals and the neighboring countries of Burundi, Congo, Rwanda and Tanzania. The Paediatric Newborn Unit has a total bed occupancy of 2.5 above capacity and on average 2000 neonates are admitted annually. The maternity ward has a bed occupation of 45 Beds with an estimated 10,000. babies born annually. Using a study done on Newborn Care Practice and Associated Factors among Mothers of One-Month-Old Infants in Southwest Ethiopia (13), 370 participants taking care of children from immunization clinic, maternity ward and newborn unit participated in the study. Ethical approvals were obtained from the Mbarara University of science and Technology Institutional review board was obtained before commencement of the study (REC No. MUST -2022-600). Written informed consent was obtained from all participants before they could participate in the study. Parents and care givers of very sick babies, whose children had died, or were sick or attending to sick mothers were excluded from the study until they were stable. We used a pre tested questionnaire to collect data on demographics, essential newborn care knowledge and practices from parents and care givers of children from birth to 6 weeks who were attending the neonatal unit and clinic, postnatal ward and the maternal and child health clinic at the referral hospital. Demographic data of the care givers, and information on their knowledge and practices about the essential care of the new born (ECNB), age, education status, income, parity, ANC attendance, family status, birth support partner was collected. New born care knowledge practices including breastfeeding (when to initiate breast feeding, frequency, prelacteal feeds, colostrum feeding), cord care, immunization, management of colic, recognition of danger signs (fever, convulsions, bleeding and jaundice among others), cultural beliefs, false teeth extraction, child scarification, use of herbs were assessed as percentages and proportions. Factors that contribute to the knowledge and practice in new – born care were analyzed using Pearson’s chi squares.

### Results

Three hundred and seventy (370) care takers were studied and majority of these were the birth mothers of the children at 86%. The mean age of the of the caretakers was 26 SD 6 years and majority of the participant were in the age range of 21-35 years at 75%. The predominant family structure was nuclear at 77.6%. Only 6.5% of the mothers were living single and not in a stable relationship. Majority of the care givers had attained only primary school education at 38.1 % while 7.6% had not received any formal education. About 73% of the caregivers were informally employed and 15% unemployed. Farming contributed 42% of the informal employment and 30.8% of the total employment of the respondents. Monthly income was mostly low with 74.6% earning below the poverty line at less than 210,000/= Uganda shillings. Maternal parity was predominantly more than 4 children 37.6%) closely followed by the prime gravida at 35.7%. Mothers had a high antenatal care attendance of 97.6% and 89% had a birth companion during the birth process. Cesarean section mode of delivery was high at 55.7% and 96.2% of the deliveries were at a health facility (Table 1)

**Table 1:** Social demographic characteristics of the participants.

| Variable | Frequency | Percentage |
| --- | --- | --- |
| **Marital status** |  |  |
| Single | 24 | 6.49 |
| Married (in a relationship) | 346 | 93.51 |
| **Care givers' age (years)** |  |  |
| $\leq 20$ | 65 | 17.57 |
| $>20 - \leq 35$ | 274 | 74.05 |
| $>35$ | 31 | 8.38 |
| **Education level** |  |  |
| None | 28 | 7.57 |
| Primary | 141 | 38.1 |
| High school | 137 | 37.03 |
| Tertiary | 64 | 17.30 |
| **Occupation** |  |  |
| Formal | 44 | 11.89 |
| Informal | 270 | 72.97 |
| None | 56 | 15.14 |
| **Income (Uganda Shillings)** |  |  |
| $<210,000$ | 276 | 74.59 |
| $>210,000 - 500,000$ | 73 | 19.73 |
| $> 500,000$ | 21 | 5.68 |
| **Mother's Parity** |  |  |
| Prime Gravida | 132 | 35.68 |
| 2-4 | 99 | 26.76 |
| $>4$ | 139 | 37.57 |
| **ANC attendance** |  |  |
| Yes | 361 | 97.57 |
| No | 9 | 2.43 |
| **Birth companion** |  |  |
| Yes | 330 | 89.19 |
| No | 40 | 10.81 |
| **Mode of delivery** |  |  |
| Cesarean section | 206 | 55.68 |
| Spontaneous Vaginal delivery | 164 | 44.32 |
| **Place of delivery** |  |  |
| Health facility | 356 | 96.22 |
| Home delivery | 14 | 3.78 |
| **Family Structure** |  |  |
| Extended | 82 | 22.40 |
| Nuclear | 284 | 77.60 |

### Essential new born care

Participant had variant practices in regard to essential newborn care. Majority (84.6%) of the respondents reported to not have put anything in the babies’ eyes at birth. Among those who reported putting something in the eyes, 70% put the recommended tetracycline eye ointment, however, other things like breastmilk, water and saliva were reportedly put in the babies’ eyes at birth. In line with keeping hygiene, 76% of the mothers washed hands with soap and water before handling their babies, while 16.2% did not wash their hands at all. About 7.4% of the new borns received a bath within 24 hours of delivery. Use of herbs to bathe babies was reported by 19% of the care givers. Caregivers practiced adequate wrapping for the babies in 87% of the children. Majority of the caregivers regularly check the baby’s umbilical cord after birth. Among the care takers who checked the cord, 36.5% report to have put something on the baby’s cord and saliva was what was frequently used followed by baby oil and herbs at 14.6% and 13.9% respectively. Only 21% of the respondents reported initiation of breast feeding within 1 hour after birth and 15.4% after 24 hours of birth. Prelacteal feeds were given by 37.6% of the care givers, water being the commonest prelacteal feed followed by cow’s milk at 40.4 and 18.4% respectively. Other prelacteal feeds included formula milk, glucose and soups. Most of the children are breastfed on demand (63.4%) while the others followed a timetabled format ranging from every 4 to 6 hours. 61.6% of the babies were reported to be vaccinated. (Table 2)

**Table 2:**
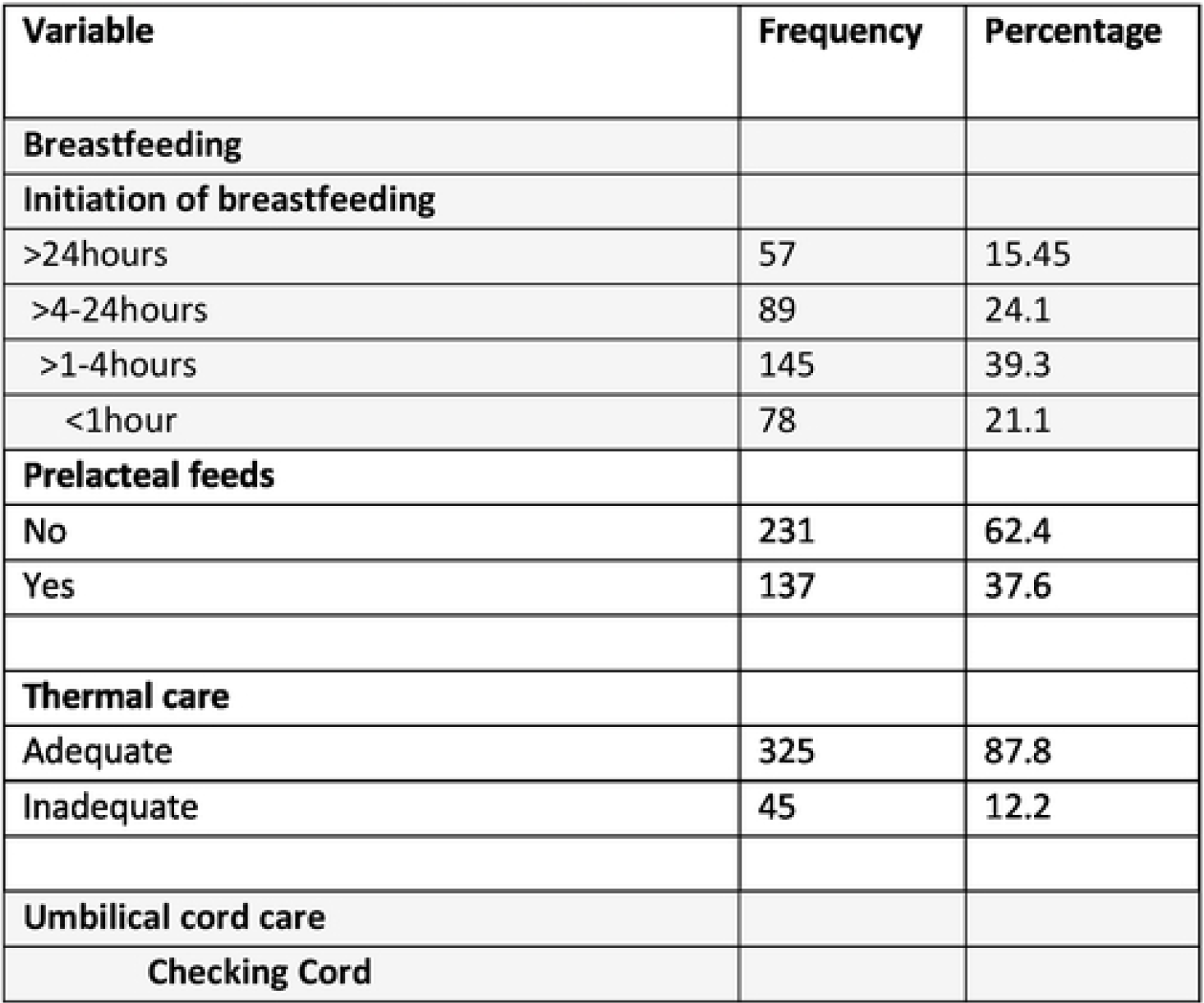
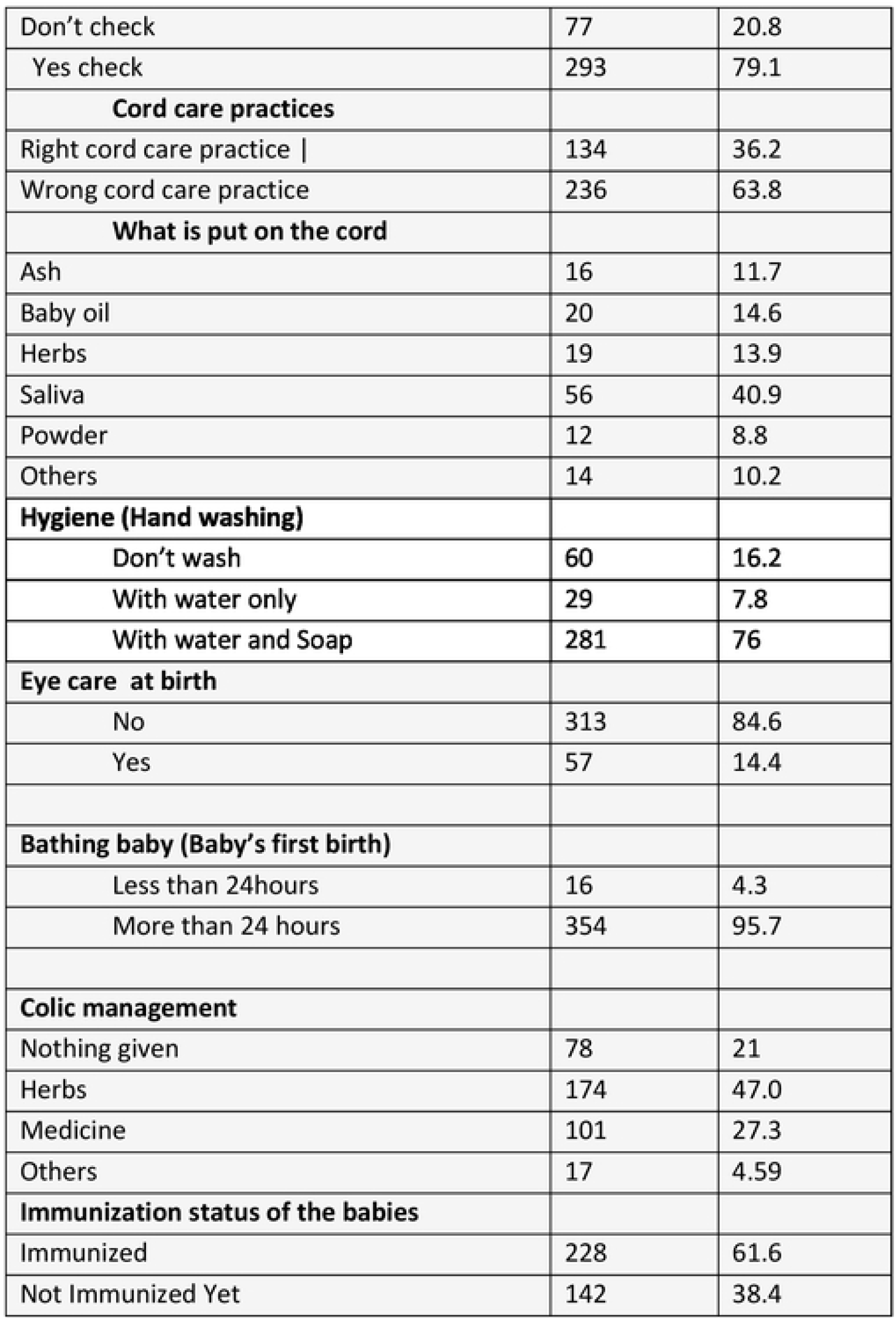
Essential newborn care practices.

| Variable | Frequency | Percentage |
| --- | --- | --- |
| **Breastfeeding** |  |  |
| **Initiation of breastfeeding** |  |  |
| >24hours | 57 | 15.45 |
| >4-24hours | 89 | 24.1 |
| >1-4hours | 145 | 39.3 |
| <1hour | 78 | 21.1 |
| **Prelacteal feeds** |  |  |
| No | 231 | 62.4 |
| Yes | 137 | 37.6 |
| **Thermal care** |  |  |
| Adequate | 325 | 87.8 |
| Inadequate | 45 | 12.2 |
| **Umbilical cord care** |  |  |
| Checking Cord |  |  |
| Don't check | 77 | 20.8 |
| Yes check | 293 | 79.1 |
| **Cord care practices** |  |  |
| Right cord care practice \| | 134 | 36.2 |
| Wrong cord care practice | 236 | 63.8 |
| **What is put on the cord** |  |  |
| Ash | 16 | 11.7 |
| Baby oil | 20 | 14.6 |
| Herbs | 19 | 13.9 |
| Saliva | 56 | 40.9 |
| Powder | 12 | 8.8 |
| Others | 14 | 10.2 |
| **Hygiene (Hand washing)** |  |  |
| Don't wash | 60 | 16.2 |
| With water only | 29 | 7.8 |
| With water and Soap | 281 | 76 |
| **Eye care at birth** |  |  |
| No | 313 | 84.6 |
| Yes | 57 | 14.4 |
| **Bathing baby (Baby's first birth)** |  |  |
| Less than 24hours | 16 | 4.3 |
| More than 24 hours | 354 | 95.7 |
| **Colic management** |  |  |
| Nothing given | 78 | 21 |
| Herbs | 174 | 47.0 |
| Medicine | 101 | 27.3 |
| Others | 17 | 4.59 |
| **Immunization status of the babies** |  |  |
| Immunized | 228 | 61.6 |
| Not Immunized Yet | 142 | 38.4 |

About 79% of the respondents reported giving something for the management of colic among babies. Herbs were the commonest remedy given (47%) followed by medicines prescribed or bought at a pharmacy (27.3%).

There was poor knowledge in identification of danger signs in the newborn by the caregivers (Figure 1). Fever was the most common identified danger sign followed by inability to breast feed in a newborn baby.

**Fig 1:**
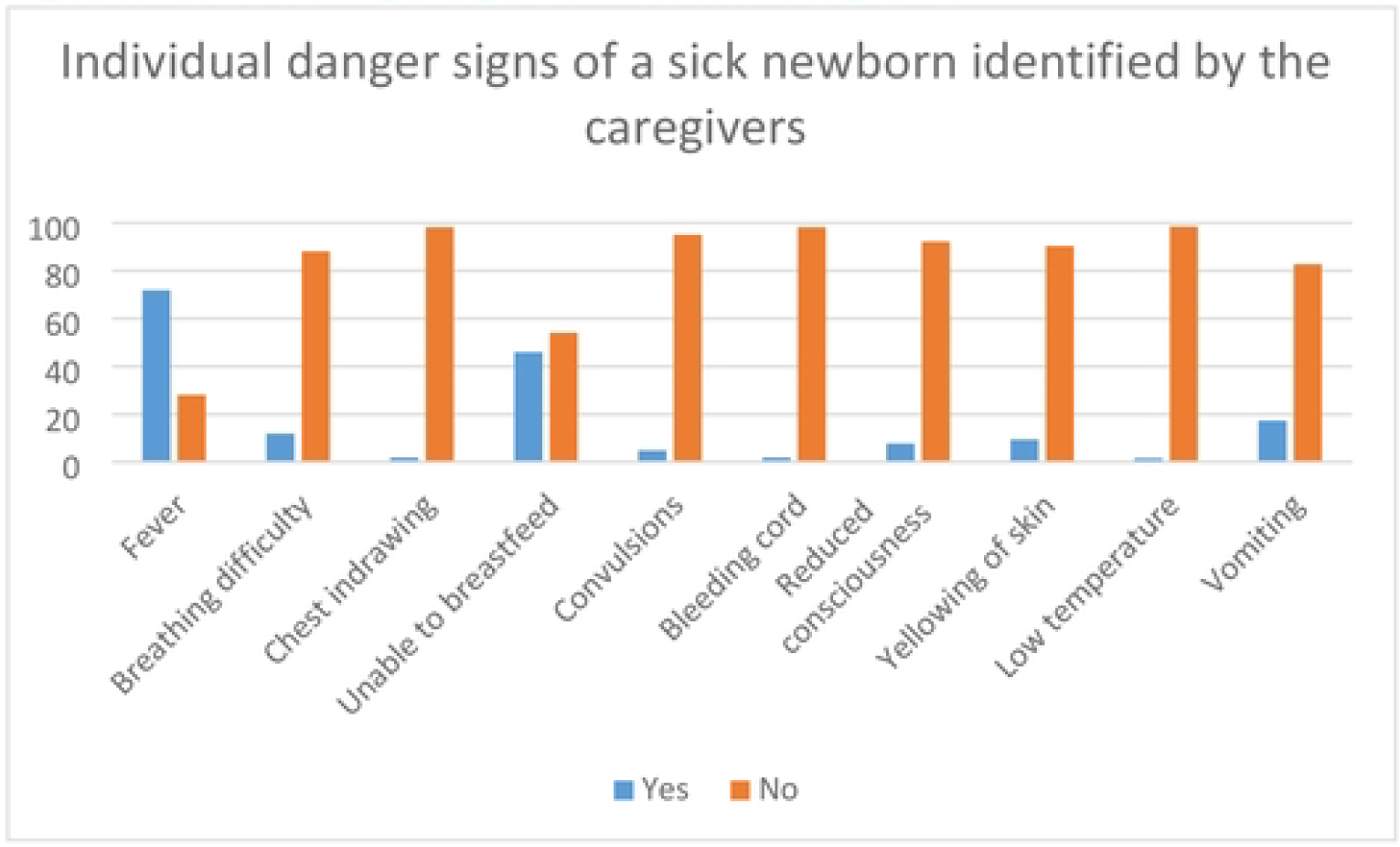
Individual danger sign score of the caregivers

Majority of the respondents had below average knowledge about danger signs in the newborn where 63% scored below 60% (Figure 3). Mean score for knowledge about danger sign was 44% SD 21% (Fig 2). Despite the low score, most respondents believed they would take a sick baby to health worker for help at 86%.

**Fig 2:**
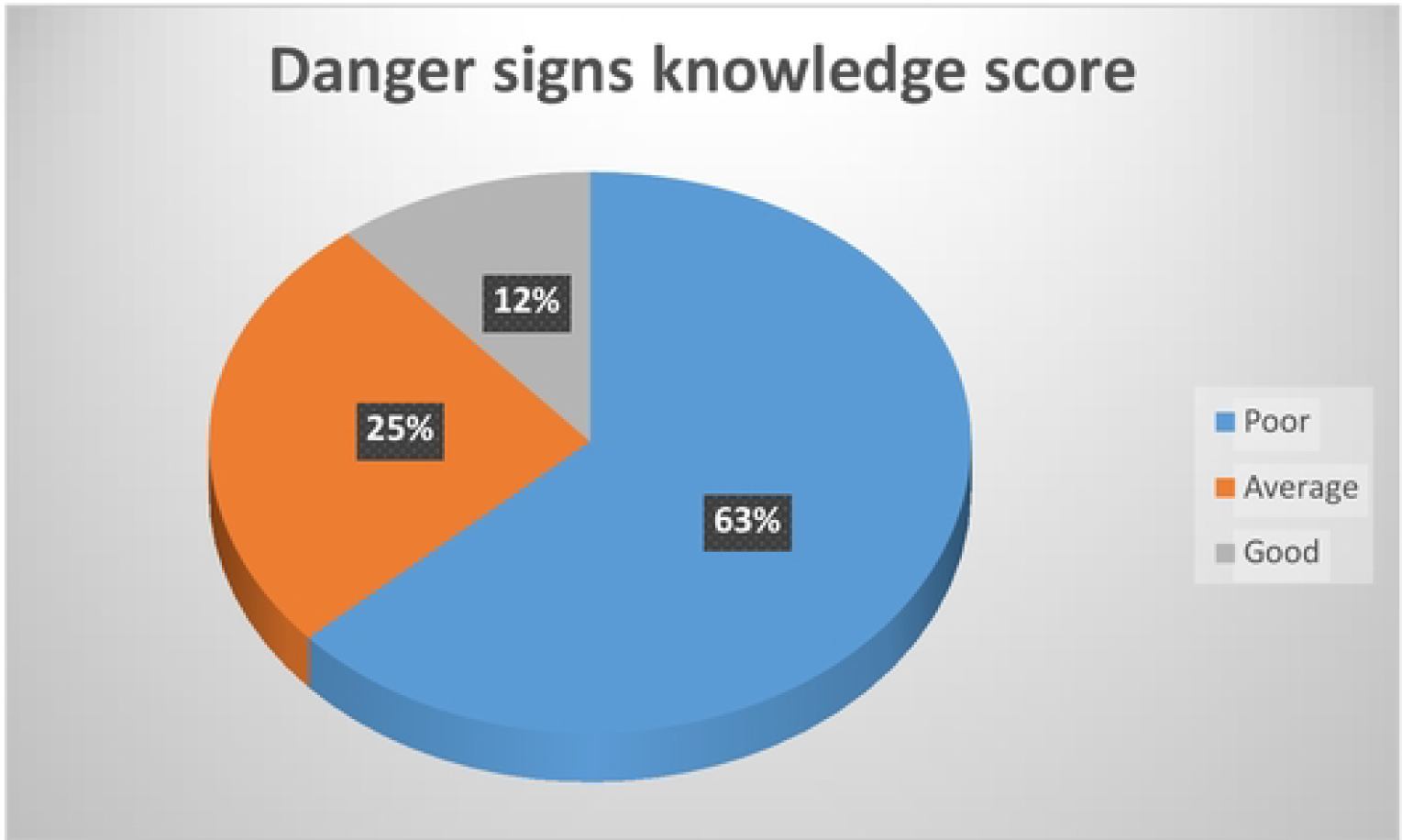
Overall danger sign knowledge score among participants

### Factors associated with newborn danger sign knowledge among caregivers

Caretaker’s age and relationship of the caretaker with the child were found to have a significant relationship knowledge of danger signs in the newborn baby at a P value of 0.019 and 0.009 respectively in a bivariate analysis (Table 3)

**Table 3:**
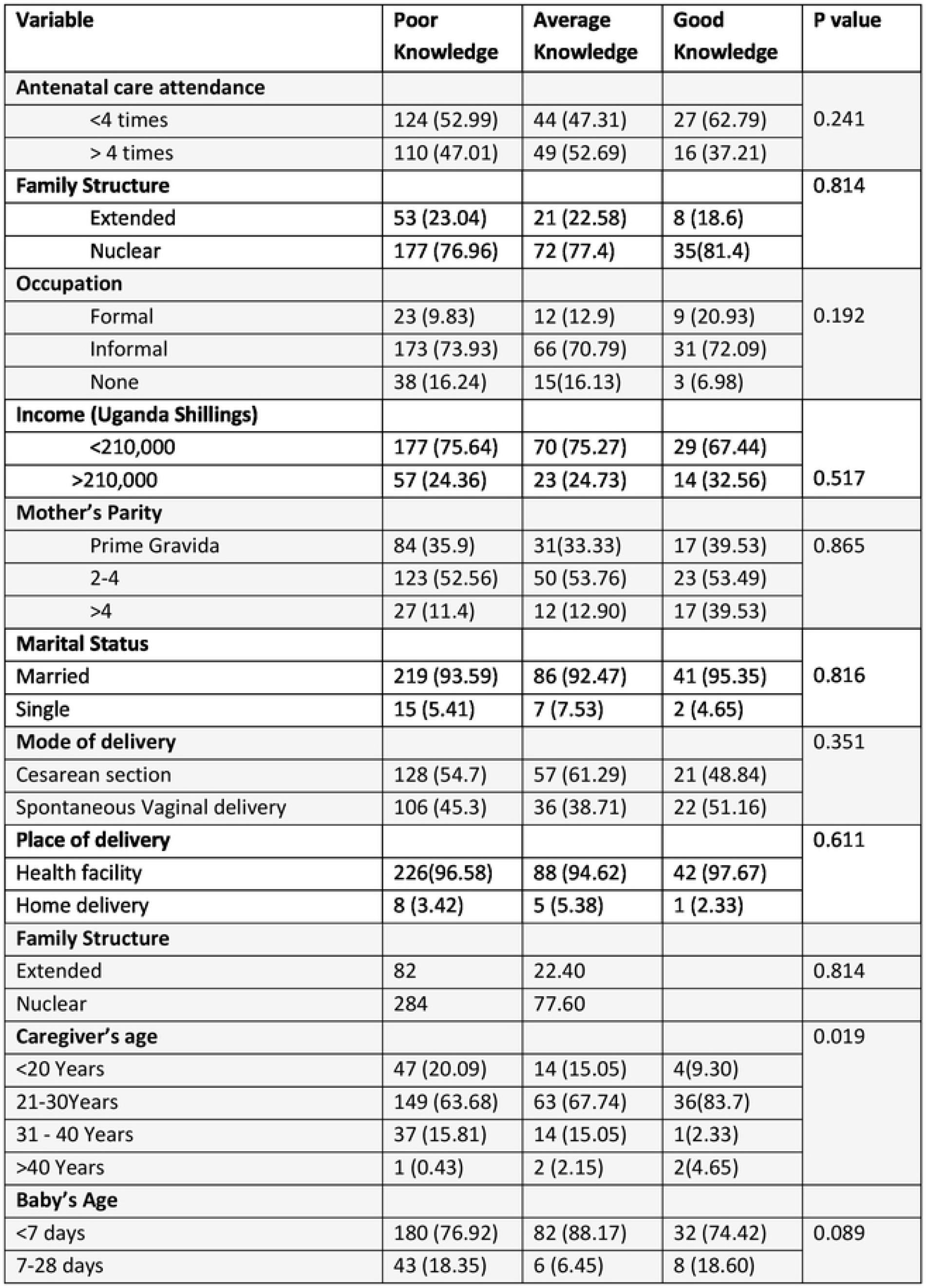
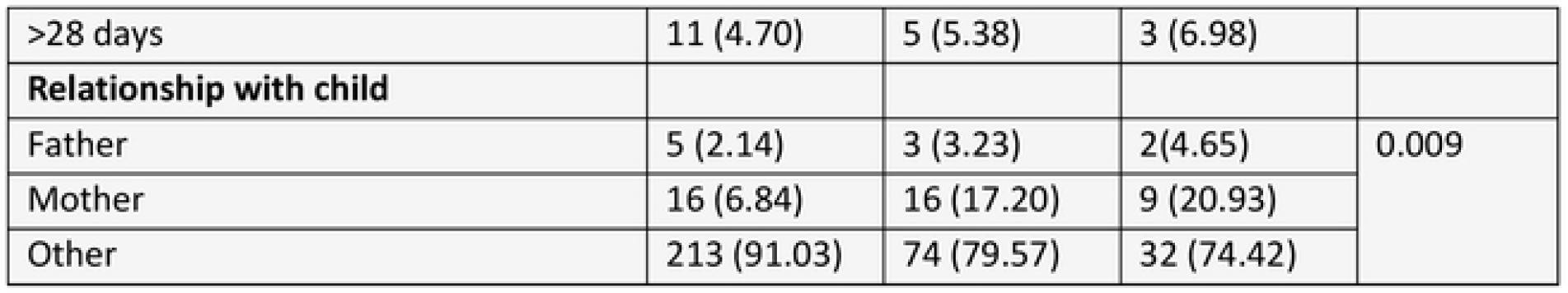
Factors associated with newborn danger sign knowledge among caregivers.

## Discussion

This study set out to evaluate newborn care knowledge and practices and the associated factors among care givers of new-born babies at a regional referral hospital in South western Uganda and we found that participants had variant levels of knowledge in the various aspects of essential new born practices and low level of knowledge of the danger signs in a new born.

Health facility deliveries were high in this study at 96.2% which is similar to what was found in Southwestern Uganda, with health facility deliveries up to 90% in the same region (14). In Tanzania and Angola, skilled birth attendance is lower at 64% and 50.7% respectively(15, 16). Antenatal care attendance was equally high at 97.57% which is similar to a study done in Angola where ANC attendance was 96.8% (15). There is a positive association of health facility delivery with antenatal care attendance(17). Furthermore, our study was a hospital study and patients were more likely to come back to a health center if they had delivered from one for different health services like immunization and health care for their children.

WHO recommends essential newborn care including early initiation of breast feeding, keeping babies warm, recognition of neonatal danger signs and cord care, among others, as crucial in new born survival(10). Contrary to WHO recommendations to initiate breastfeeding within an hour of birth, only 21.1% initiated breastfeeding within 1 hour after birth, and 15.45% of the newborns initiated breastfeeding after 24 hours in this study. This is much lower than what was found in a study done in India(18). This disparity is explained by the fact that India has well established breast feeding programs which are absent in the current study setting. Furthermore, prelacteal feeds were administered in 37.6% of the newborn children. This is lower than 64.7% found in Pakistan(19) probably because most of the birth in our study occurred in a health facility. Although lower than in Pakistan, this proportion of prelacteal feeds was higher than pooled proportion in East Africa(20). The east African study included children that had been born 5 years earlier and this could have brought about recall bias compared to the children in our study that were less than six weeks of age.

Majority of the respondents purposefully checked their baby’s cord, however reported wrong cord care practices with majority (63.8%) applying substances such as Ash, saliva, herbs and baby oil, which practice is contrary to standard and recommended care and puts newborn babies at risk of dying(21). This however is similar to what has been found in Ghana (64.3%) (22) and Ethiopia with malpractice in cord care up to 66.9%(13).

In this study, care givers practiced adequate thermal care (87.8%) practicing wrapping the baby in multiple layer to keep the baby warm. This is lower than 60.9% found in a community study done in India(23). Ours was a hospital based study and the respondents could have had health education that promoted adequate thermal care. However, 24% of the newborns were given a bath within 24 hours of delivery. This finding is lower than what was found in a study done in Ethiopia where 32.5% of mothers had practiced early bathing of the newborn(24). In Bangladesh, only 10.2 % of mothers bathed their babies by the first day of cultures. These lower values may be attributed to differences in culture and beliefs surrounding birth between Africa and Asia.

About 38.4% of the children were not immunized up to date despite the mean age being 7 days. This may be corresponding with the level of immunization coverage in the region, but also since some of the children were in hospital, they may have differed immunization until after discharge when the children were better. In addition, the facility has one point of immunization without finding patients on maternity or paediatrics wards. A study done in India found similarly low vaccination uptake of 47.4% for children less than 42 days (25) which may be attributed to access and attitudes of the caregivers to vaccination services.

Proper hand washing with soap and water before handling the baby was not practiced in 24% of the respondents. This may be due to cultural beliefs and lack of knowledge in regards to hygiene, but also lack of access to proper hand washing facilities at their homes and the newborn care facilities as identified in Nigeria and parts of Asia (26, 27). In addition, majority of the participants in the study were below poverty line which may pose a challenge in accessing basic needs like soap and clean water.

A big number of respondents were ignorant of eye care for the new borns with 84.6% unaware of the need to put any medication in the babies’ eyes at birth. This is similar to studies done in India(28) which shares similar social and cultural beliefs to Uganda

Regarding colic and its management, majority of the respondents (79%) had wrong practices that involved administration of herbs and over the counter medicines to alleviate the pain. This may be explained by lack of knowledge about appropriate measure and the caregivers’ sense of helplessness when the child is in pain (29)

Care giver’s knowledge about new born danger signs was low with a mean score of 44% which is similar to what has been found in Saudi Arabia and Ethiopia at 37% and 48.3% respectively (30, 31). The low knowledge may be due to lack of targeted education programs for newborn caregivers focusing on the sick newborn. The commonly recognized danger sign was fever at 71% of the respondents. Despite the relatively low knowledge about danger signs of a sick new born, majority of the respondents reported seeking care from a health worker and only 13.1 % seeking care and attention from other alternatives like relatives, spiritual leaders and herbalists.

In conclusion, this study reveals that there is various malpractice in the essential new born care and low level of knowledge of danger sign in a new born among caregivers of newborn babies. There is an increased health center deliveries and antenatal care attendance among the respondents. This improved health services utilization could be used as an avenue to increase awareness about the malpractices in essential newborn care and also increase awareness about the danger signs in a newborn among caregivers of newborn children.

The strength of this study is that it highlighted the gaps in practice of essential newborn care by caregivers and reduced awareness among both parents and caregivers of newborn babies. This information could be used as baseline data in planning knowledge and practice enhancement programs for mothers and caregivers of new born babies.

The limitation of this study was that it was done in an institution and may not represent the caregivers and parents in the community. A community study could help evaluate any differences in knowledge and practice in care of the newborn.

## Data Availability

Data cannot be shared publicly because of condidentiality policy. Data are available from the corresponding author (contact via) for researchers who meet the criteria for access to confidential data.

## Declarations

Ethics approval and consent to participate

This study was approved by the Mbarara University of Science and Technology Research Ethics Committee (REC No. MUST -2022-600) and administrative clearance was obtained, Mbarara Regional Referral Hospital, prior to conducting the study. Confidentiality of the study participants was ensured by using unique identifiers and the charts were kept in a lockable cupboard which was accessible by only the study team

## Availability of data and materials

The datasets generated and analyzed for this study are available from the corresponding author, upon request.

## Consent for publication

All authors have consented to the publication of this research work

## Authors’ contributions

DN, SK and EK contributed to the conception and design of the study. DN and EK performed formal data analysis. DN, SK, JN, EN, OB, LK and JK contributed to drafting the manuscript and contributed to study implementation and data acquisition. DN and EK critically reviewed and revised the manuscript for key content. DN prepared the final manuscript. All authors read and approved the final manuscript.

